# Vaccine serologic responses among transplant patients associate with COVID-19 infection and T peripheral helper cells

**DOI:** 10.1101/2021.07.11.21260338

**Authors:** Jacob E. Lemieux, Amy Li, Matteo Gentili, Cory A. Perugino, Zoe F. Weiss, Kathryn Bowman, Pierre Ankomah, Hang Liu, Gregory D. Lewis, Natasha Bitar, Taryn Lipiner, Nir Hacohen, Shiv S. Pillai, Marcia B. Goldberg

**Author notes:** Corresponding authors: Division of Infectious Diseases, Massachusetts General Hospital, 55 Fruit St., Boston, MA, 02114, USA, and.

## Abstract

**Background:** Therapeutically immunosuppressed transplant recipients exhibit attenuated responses to COVID-19 vaccines. To better understand the immune alterations that determined poor vaccine response, we correlated quantities of circulating T and B cell subsets at baseline with longitudinal serologic responses to SARS-CoV-2 mRNA vaccination in heart and lung transplant recipients.

**Methods:** Samples at baseline and at approximately 8 and 30 days after each vaccine dose for 22 heart and lung transplant recipients with no history of COVID-19, four heart and lung transplant recipients with prior COVID-19 infection, and 12 healthy controls undergoing vaccination were analyzed. Anti-spike protein receptor binding domain (RBD) IgG and pseudovirus neutralization activity were measured. Proportions of B and T cell subsets at baseline were comprehensively quantitated.

**Results:** At 8-30 days post vaccination, healthy controls displayed robust anti-RBD IgG responses, whereas heart and lung transplant recipients showed minimally increased responses. A parallel absence of activity was observed in pseudovirus neutralization. In contrast, three of four (75%) transplant recipients with prior COVID-19 infection displayed robust responses at levels comparable to controls. Baseline levels of activated PD-1^+^ HLA-DR^+^ CXCR5^-^ CD4^+^ T cells (also known as T peripheral helper [T_PH_] cells) and CD4+ T cells strongly predicted the ability to mount a response.

**Conclusions:** Immunosuppressed patients have defective vaccine responses but can be induced to generate neutralizing antibodies after SARS-CoV-2 infection. Strong correlations of vaccine responsiveness with baseline T_PH_ and CD4^+^ T cell numbers highlights a role for T helper activity in B cell differentiation into antibody secreting cells during vaccine response.

## Introduction

For immunocompetent individuals, SARS-CoV-2 vaccination is a highly effective preventative approach for protection from COVID-19 (1–3). However, because Phase 3 trials excluded immunosuppressed individuals, the approach to and determinants of induction of protective immunity against SARS-CoV-2 among transplant recipients and other severely immunosuppressed individuals is less clear.

Heart and lung transplant recipients require a higher degree of immunosuppression to prevent graft rejection than other solid organ transplant recipients. Their regimens typically include a calcineurin inhibitor (tacrolimus more often than cyclosporine), a DNA synthesis inhibitor (mycophenolate more often than azathioprine) and/or a mammalian target of rapamycin (mTOR) inhibitor (sirolimus), and low-dose glucocorticoids. Whereas both DNA synthesis inhibitors and calcineurin inhibitors block T cell expansion, tacrolimus may particularly impact follicular helper T cells (4), which are critical for the affinity maturation and isotype switching of an effective antibody response. Loss of mTOR activity not only blunts T and B cell proliferation, but also impairs the development of CD8+ T cell memory (5), skews helper T cell differentiation toward regulatory T cells (6, 7), and disrupts B cell progression through germinal center reactions, including class switching and somatic hypermutation (8–10). Furthermore, glucocorticoids interfere with activities of virtually all immune cells.

Previous to COVID-19, several reports demonstrated attenuated or delayed serologic responses to various vaccines among severely immunosuppressed hosts (11–15). Vaccine efficacy in this patient population may depend in part on the nature of the vaccine and is sometimes improved by booster vaccination (16). Published data on antibody responses to SARS-CoV-2 mRNA vaccination in transplant recipients indicate major impairments in vaccine-induced humoral and cellular immunity (16–25). Whether transplant recipients are uniquely vulnerable to more transmissible viral strains with spike protein variants, which are rising in prevalence globally, remains unclear. Hence, there is an urgency to determine why transplant recipients do not respond to standard SARS-CoV-2 mRNA vaccinations and to identify solutions to protect this vulnerable patient population.

Here, we quantitatively assessed the antibody response to SARS-CoV-2 vaccination among heart and lung transplant recipients, including neutralization activity to emerging variants of concern, extending recent findings from other groups (17, 22–25). Further, to identify cellular correlates of adaptive immune determinants of vaccine responsiveness, we quantified subsets of CD4^+^ T helper cells, CD8+ T cells, and B cells at baseline and correlated these with levels of serum IgG.

## Results

### Serum IgG responses among vaccinated heart and lung transplant recipients were markedly diminished compared to those of healthy controls

Among vaccinated heart and lung transplant recipients with no evidence of prior or intercurrent (between vaccine doses) COVID-19 infection (N=16 heart and N=6 lung transplant recipients), levels of IgG recognizing the SARS-CoV-2 RBD were markedly lower than those of healthy controls (Fig. 1). At 8 and 30 days after the second vaccine dose, IgG anti-RBD levels were 22-fold lower (p = 2.9 × 10^−7^; Wilcoxon rank-sum test) and 20-fold lower (p = 6.7 × 10^−4^; Wilcoxon rank-sum test), respectively, among transplant recipients compared to those of healthy controls.

**Figure 1:**
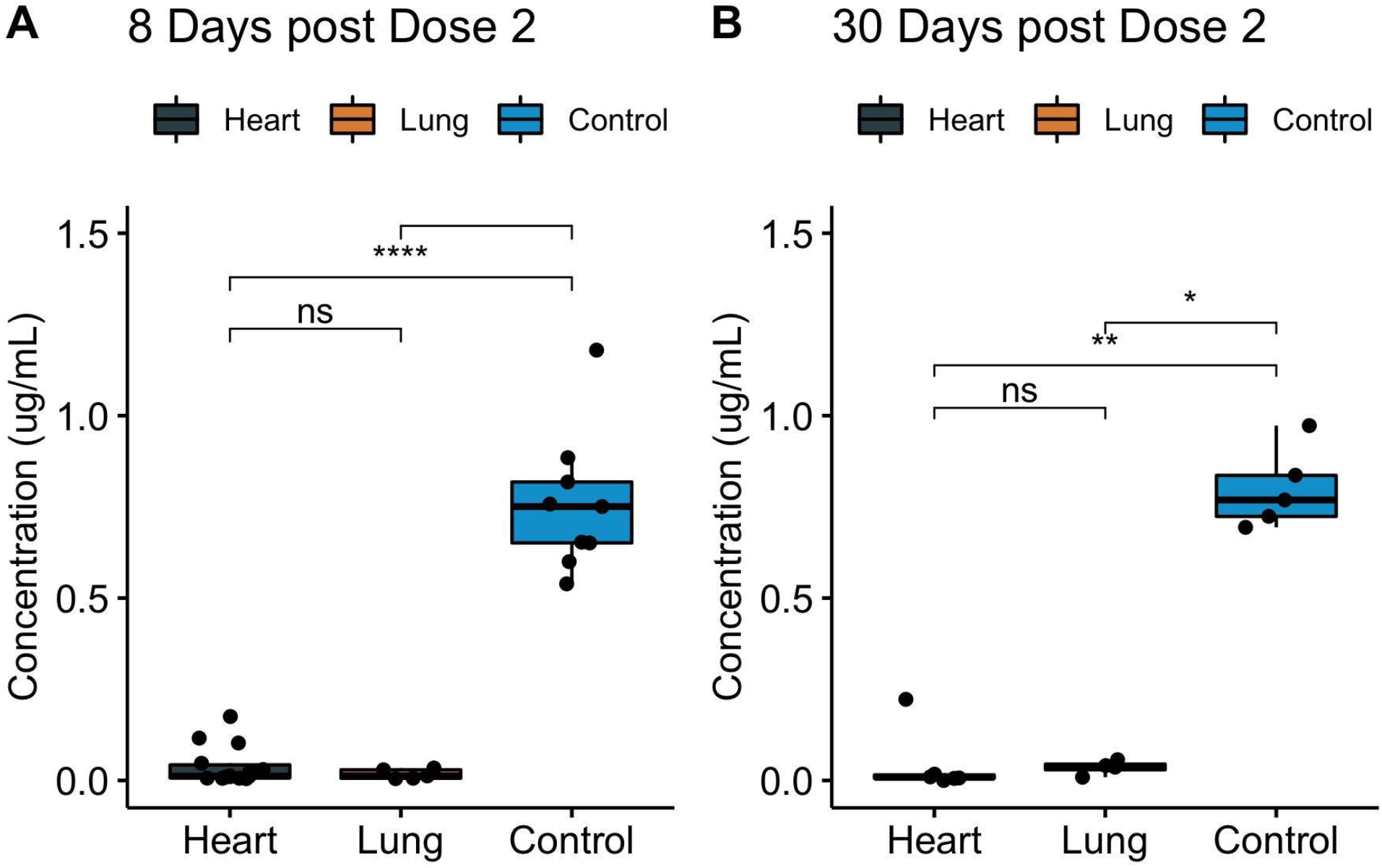
Levels of serum IgG to SARS-CoV-2 spike protein RBD among fully vaccinated COVID-19-naive heart and lung transplant recipients. Serum IgG binding to RBD, as measured by ELISA, at 8 days (**A**) or 30 days (**B**) after full vaccination, displayed by transplanted organ type. Data for the transplant recipients who had prior COVID-19 infection are excluded. Boxes, 25^th^, 50^th^, and 75^th^ percentile; whiskers, smallest and largest values in dataset up to 1.5x interquartile range. All data points are shown. ^*^, p < 0.05; ^**^, p < 0.01; ^****^, p < 0.0001.

### Neutralization activity of plasma from vaccinated heart and lung transplant recipients were markedly diminished compared to that of healthy controls

Neutralization activity to SARS-CoV-2 pseudovirus among vaccinated heart and lung transplant recipients largely paralleled IgG levels. At 8 days after the second vaccine dose, mean neutralizing antibody activity of transplant recipients was 12.2-fold less than that of healthy controls (p = 2.9 × 10^−7^, Wilcoxon rank-sum test, Fig. 2A), and at 30 days after the second vaccine dose, neutralizing antibody activity of transplant recipients was 12.0-fold less than that of healthy controls (Wilcoxon rank-sum test, p = 0.001, Fig. 2B).

**Figure 2:**
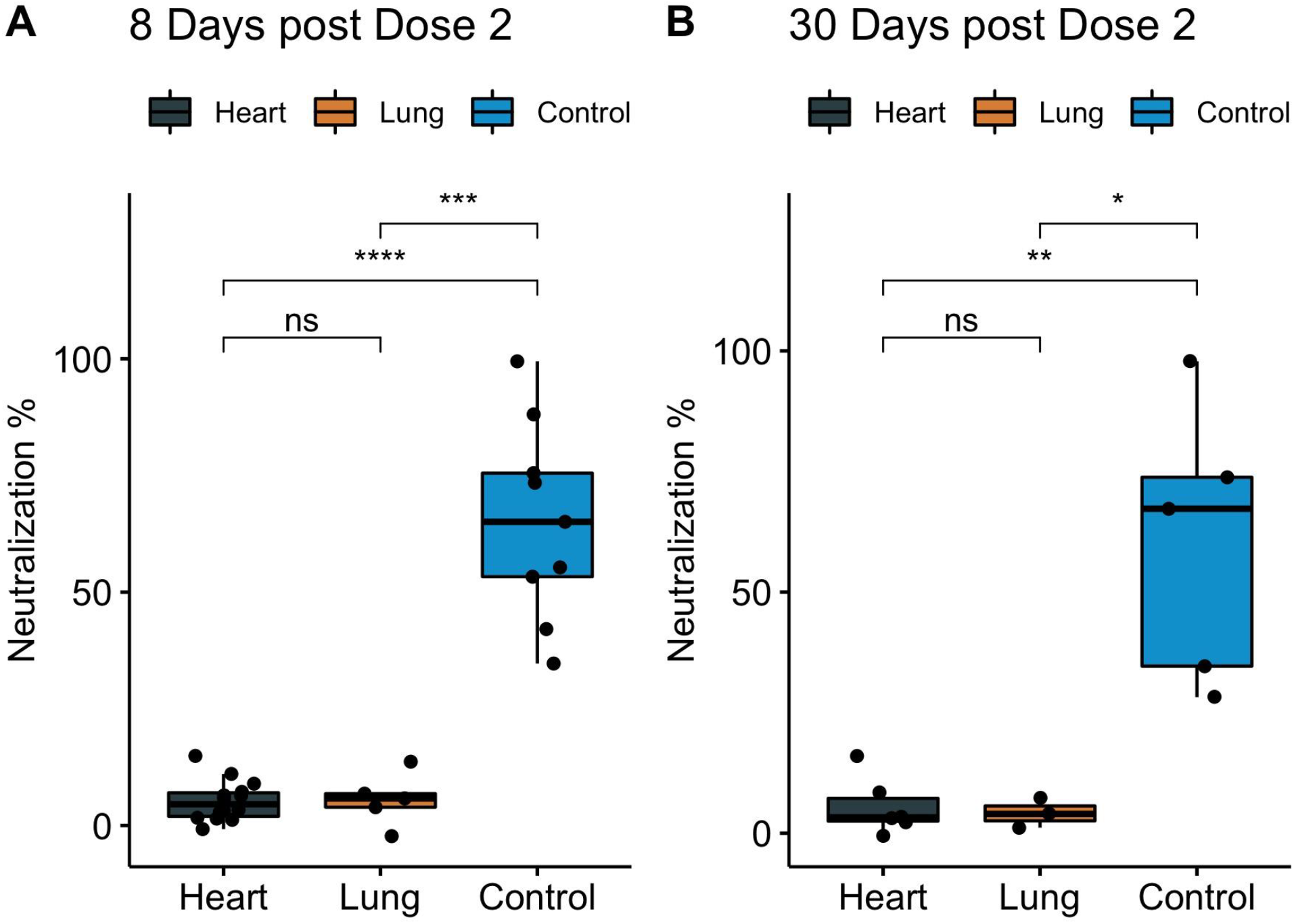
Plasma pseudovirus neutralization activity among fully vaccinated COVID-19-naive heart and lung transplant recipients. SARS-CoV-2 neutralization, as measured by pseudovirus neutralization assay, at 8 days (**A**) or 30 days (**B**) after full vaccination, displayed by transplanted organ type. Data for the transplant recipients who had prior COVID-19 infection are excluded. Boxes, 25^th^, 50^th^, and 75^th^ percentiles; whiskers, smallest and largest values in dataset up to 1.5x interquartile range. All data points are shown. ^*^, p < 0.05; ^**^, p < 0.01; ^***^, p < 0.001; ^****^, p < 0.0001.

Similar to previously published results (26–31), among healthy controls, neutralizing activity to variant pseudoviruses containing the E484K/N501Y/D614G mutations present in the beta (B.1.351) and gamma (P.1) variants (originally identified in South Africa and Brazil, respectively) were reduced (Fig. S1C). Among control patients at later time points (1d pre dose 2 through 68d post dose 2), neutralization against E484K/N501Y/D614G pseudovirus was 22% lower than against D614G pseudovirus (p = 6.6 × 10^−6^, paired Wilcoxon test) and 24% lower than against N501Y/D614G pseudovirus (p = 3.8 × 10^−5^, paired Wilcoxon test) (Fig. S1C), representative of the alpha (B.1.1.7) variant originally identified in the United Kingdom; the difference in neutralization between D614G and N501Y/D614G pseudoviruses was not significant. Among both transplant recipients and healthy controls, neutralizing antibody titers correlated strongly with levels of anti-RBD IgG, although this correlation decreased with additional variant mutations in the RBD (Fig. S2).

### Time course of response

The differences in responses between the transplant recipients and the healthy controls were evident as early as 20 days after the first vaccine dose (1 day prior to the second vaccine dose), when all but two healthy controls yet none of the transplant recipients displayed increases in IgG above background (Figs. 3, S3, and S4). Following the second dose of vaccine, IgG levels and neutralization activity progressively increased in two COVID-19-uninfected transplant recipients and all of the controls (Figs. 3 and S4). Increases in neutralization activity correlated highly with the increases in IgG levels (for D614G pseudovirus and IgG levels, *R*=0.88, Fig. S2A). The increases in these two transplant recipients were delayed compared to the controls; whether they will continue to increase at later times will be determined with additional follow-up. Consistent with prior publications in which lower immunosuppression is associated with improved serologic responses (17, 22–24), these two transplant recipients were on levels of immunosuppression at the lower end of our cohort; one was not on a DNA synthesis inhibitor, and both were on only 5 mg daily of prednisone.

**Figure 3:**
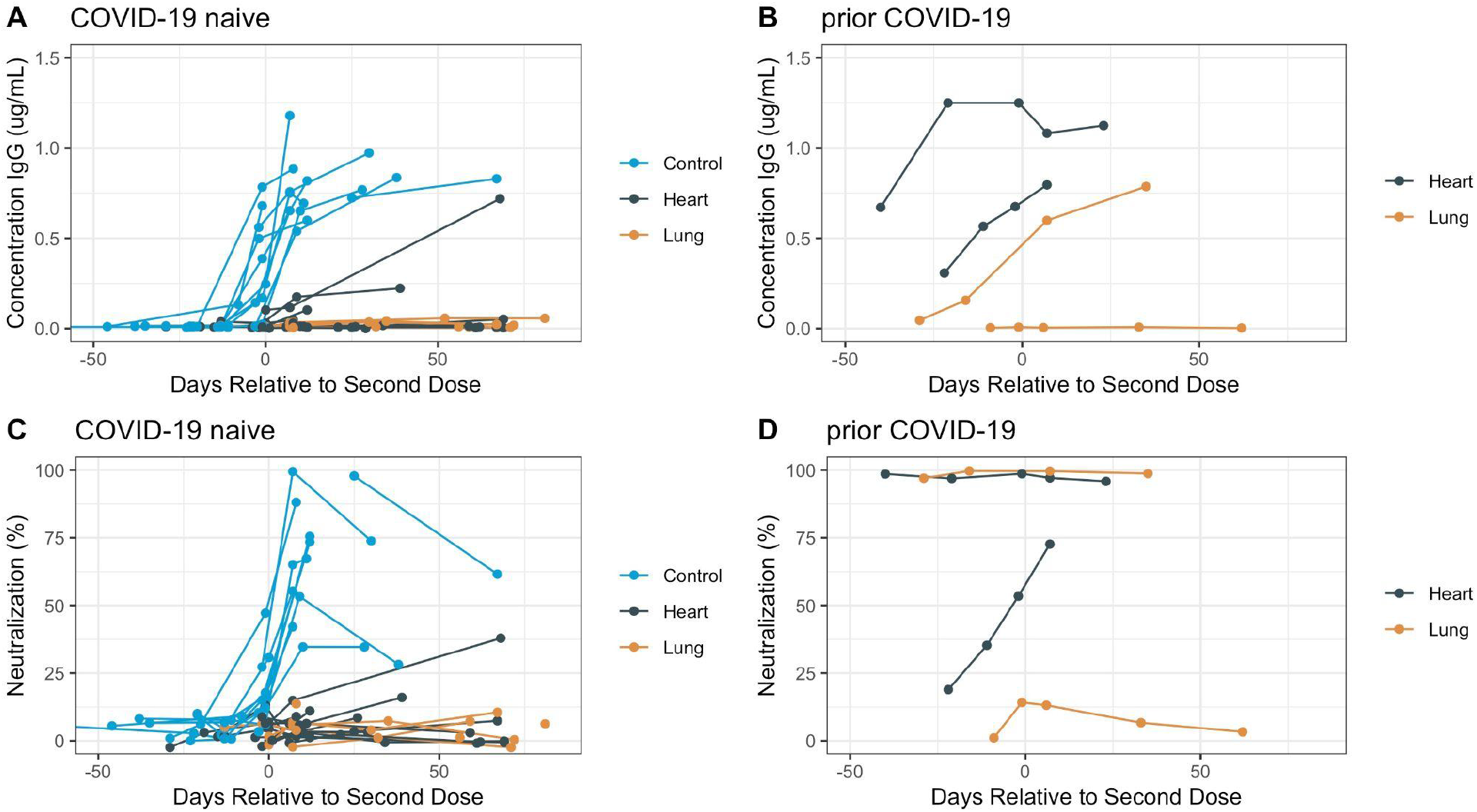
Antibody trajectories over time in individual subjects. (**A**-**B**) IgG binding to SARS-CoV-2 spike protein RBD over time, as measured by IgG ELISA, in COVID-19 naive subjects (**A**) or in subjects with prior COVID-19 infection (**B**) displayed by transplanted organ type. Neutralization activity over time in COVID-19 naive subjects (**C**) or in subjects with prior COVID-19 infection (**D**). All data points are shown.

### Prior or intercurrent infection with COVID-19 is associated with strong serological responses and neutralizing activity

Three transplant recipients had COVID-19 prior to vaccination, and one had COVID-19 between the first and second vaccine dose. Three of these four transplant recipients displayed IgG levels and neutralization activity were comparable to those of healthy controls (Fig. 3B, 3D). For the three of these individuals who displayed elevated IgG levels and neutralization activity, vaccination *per se* was associated with a boost in IgG levels, and for one, a boost in neutralization activity; for the other two who responded, it was not possible to determine whether neutralization activity was boosted, as it was >95% at baseline (Fig. 3D). The proportion of subjects who responded to vaccination was significantly different among transplant patients with prior COVID-19 (p = 0.018, Fisher’s exact test). Proximity to transplantation and the higher doses of immunosuppressive medications generally administered around transplantation may have contributed to the absence of response in the COVID-19-infected non-responder, as this individual developed infection <1 year after transplantation, whereas the other three COVID-19-infected transplant recipients developed infection 2-5 years (N=2) and >15 years (N=1) after transplantation. No differences in immunosuppressive regimens at the time of vaccination or the severity of COVID-19 infection between the one COVID-19-infected non-responder and the three COVID-19-infected responders were identified. Nor were any differences in immunosuppressive regimens identified between the COVID-19-infected and uninfected transplant recipients.

### Peripheral blood immune cell populations at early time points and cellular predictors of antibody response

To identify differences that may underlie the differential response to vaccination, we performed high-dimensional flow cytometry on baseline samples and quantified peripheral blood immune cell subsets at early time points. We found significant differences in peripheral blood immune cells including CD3+ lymphocytes (p = 0.04, wilcoxon test), CD4+ T lymphocytes (p = 0.002), T peripheral helper (T_PH_) cells, Tregs (p = 0.02) and DR+ T cells (p = 0.0059). The relative proportion of T_PH_/CD4+ (p = 0.0093), DR+/Tregs (p = 0.00031), and DR+/CD4+ (p = 0.04) were also decreased among transplant patients as compared to controls (Fig. 4A and S5A). In contrast, we did not observe significant differences in CD8+ T cells and CD19+ B cell populations. To identify potential cellular predictors of the antibody response to vaccination, we computed correlation statistics between baseline flow cytometry and antibody response at 8 days post second dose (Fig. S5B). Most strongly correlated with antibody response was the absolute number of T peripheral helper (T_PH_) cells (Pearson correlation coefficient, r = 0.72, p = 0.0036), and this relationship held for the proportion of T_PH_ cells among total CD4^+^ cells (r = 0.62, p = 0.02). T_PH_ cells are PD-1^+^ CXCR5^-^ HLA-DR^+^ CD4^+^ T cells and represent a subset of CD4^+^ T cells that have been shown to drive B cell differentiation into antibody secreting cells (32). A substantial yet weaker relationship was present between day 8 IgG levels and HLA DR+ CD4+ T cells (Fig. 4B).

**Figure 4:**
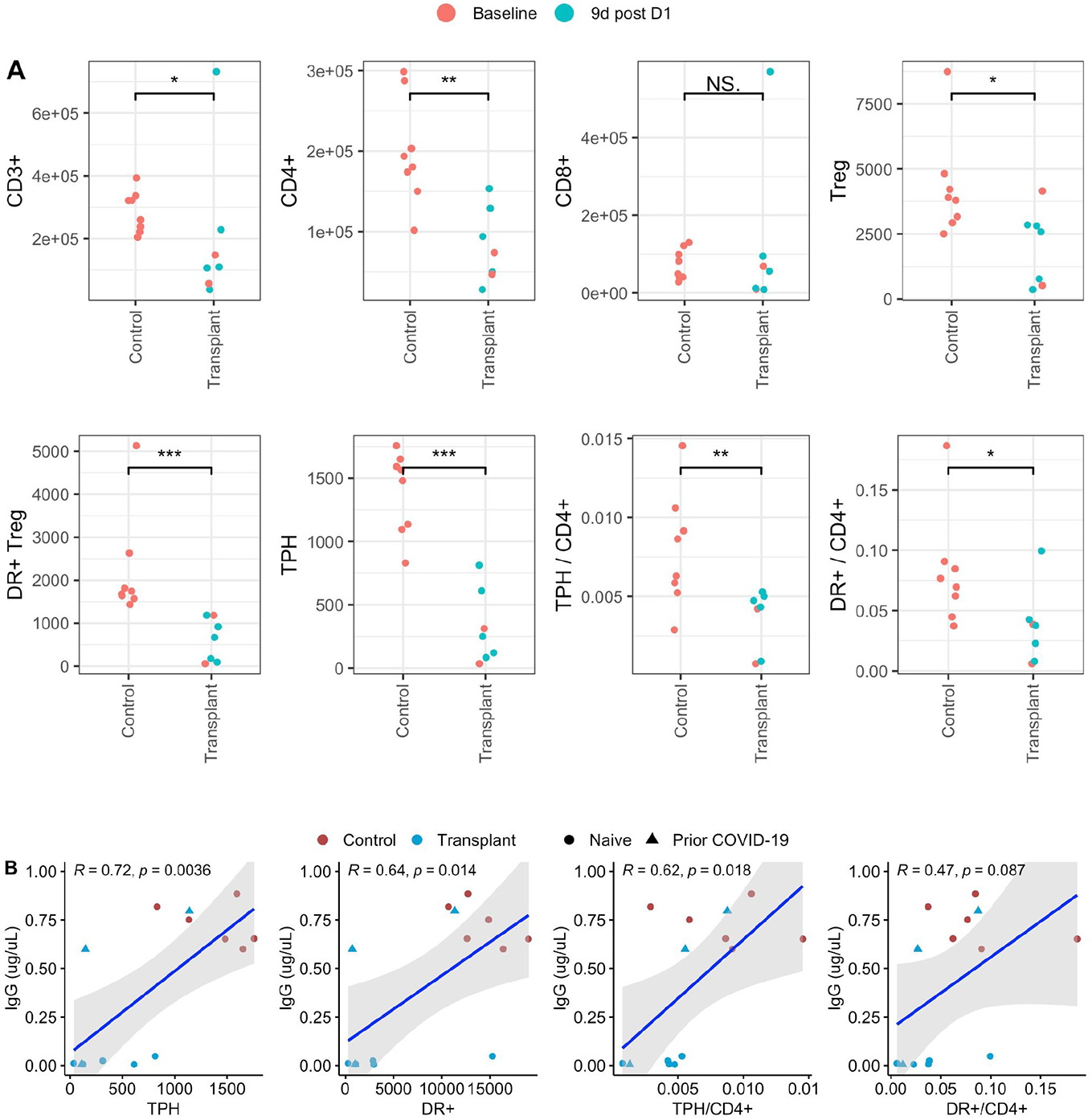
Immune cell subsets and cellular correlates of antibody response. (**A**) Quantification by flow cytometry of peripheral blood cells in COVID-19-naive individuals. (**B**) Correlation between antibody response at day 8 after complete vaccination and baseline levels of DR+ CD4 T peripheral helper (TPH) cells; DR+ CD4 T cells; DR+ CD4 T cells as a proportion of total CD4+ cells at baseline, or TPH cells as a proportion of total CD4+ cells at baseline. Least squares regressions (blue lines). Gray shading, 95% confidence interval (CI). *R*, correlation coefficient. All data points are shown. ^*^, p < 0.05; ^**^, p < 0.01; ^***^, p < 0.001; ^****^, p < 0.0001.

We observed no significant correlations between serologic responses and the baseline proportions of total effector-memory CD4^+^ T cells, regulatory T cells, HLA-DR^+^ (activated) regulatory T cells, naïve CD4^+^ T cells, or circulating T follicular helper T cells among total CD4^+^ T cells, effector-memory CD8^+^ T cells, CD8^+^ cytotoxic T cells, or CD8^+^ naïve T cells among total CD8^+^ T cells, naïve, unswitched memory, switched memory, or double-negative (IgD^-^CD27^-^) B cells among total CD19^+^ B cells.

## Discussion

In this study, we provide the first in-depth longitudinal comparison of immune responses to SARS-CoV-2 vaccination in heart and lung transplant recipients to responses to vaccination of healthy controls and of heart and lung transplant recipients with prior COVID-19 infection. In addition, we provide the first correlation of these responses to quantitative analyses of baseline T and B cell subsets. Consistent with findings of other investigators (16–25), we show that COVID-19-naive heart and lung transplant recipients have a profound reduction in neutralizing antibodies against SARS-CoV-2 spike protein and receptor binding domain (RBD)-specific immunoglobulin levels after both prime and boost vaccine doses. However, transplant recipients who had COVID-19 infection prior to vaccination displayed neutralizing antibodies to SARS-CoV-2 at baseline that increased following vaccination. Taken together, the disparate humoral immune response to SARS-CoV-2 vaccine as compared to natural infection in heart and lung transplant patients is suggestive of a failure of vaccines to induce immunologic priming in highly immunosuppressed patients, and points to the need for alternate immunization regimens for these populations.

The vast majority of participants received the two-dose BNT162b2 (Pfizer-BioNTech) mRNA vaccine (88%), with only two heart and lung transplant recipients (7.7%) receiving the two-dose mRNA-1273 (Moderna) vaccine and only one (3.8%) receiving the single-dose Ad26.COV2.S (Janssen) adenovirus-based vaccine; as a result, we were unable to assess vaccine-related outcomes and our outcomes in aggregate are largely reflective of responses to the Pfizer vaccine.

Our data are consistent with recent studies showing response rates of 0-49% in heart and lung transplant recipients receiving the BNT162b2 mRNA vaccine (22, 24, 33, 34). These studies provided evidence of lower anti-spike IgG levels in transplant recipients; we show that this is corroborated by lower neutralizing antibody titers as well. These studies also showed an association between poor response and the use of various immunosuppressive regimens, particularly those including DNA synthesis and calcineurin inhibitors. Compared with our findings, the higher response rates (up to 49%) observed in heart and lung transplant recipients in three of these studies (22, 24, 33) may reflect lower proportions of patients on combined calcineurin and DNA synthesis inhibition as well as maintenance steroids, whereas the vast majority of transplant recipients described in Havlin et al. (34) and in our cohort, who had a uniform lack of response to vaccine, are maintained on three-drug therapy.

The presence of robust neutralizing antibody titers in 3 of the 4 transplant recipients with prior COVID-19 infection in our study suggests that heavily immunosuppressed patients can nevertheless mount an effective humoral response to natural infection. This is in line with Havlin et al., who found that 85% of lung transplant recipients with SARS-CoV-2 infection developed spike-directed antibodies (34). The higher proportion of T_PH_ cells in transplant patients who had prior COVID-19 infection may suggest an expansion in this population due to recent infection, which may in turn facilitate the boost in antibody responses seen in these individuals. A corollary to this is that the uniformly low numbers and proportion of T_PH_ cells in uninfected transplant patients may underlie the inability of these individuals to mount spike-specific IgG responses. Our data indicate that mRNA vaccination may be sufficient to boost existing responses, as anti-spike antibody levels rose post-vaccination in patients with prior COVID-19 infection with similar kinetics to that of healthy controls; however, mRNA vaccination is a far weaker driver of immunologic priming of immune responses than natural infection.

The generation of long-lasting humoral immunity depends on a coordinated T and B cell response to antigen. Calcineurin inhibitors have been shown to impact antibody responses by inhibiting T follicular helper (T_FH_) cell differentiation, as well as naive B cell proliferation and plasmablast differentiation (4, 35). In our data, the substantial deficits in total and activated CD4+ in transplant patients at baseline suggests that a failure to mount appropriate T cell help for B cell differentiation results in impaired class switching and memory B cell development. A limitation of our analysis is that pre-vaccine measurements were available only for 3 transplant patients and our “baseline” populations among transplant patients were mixtures of pre-vaccine and 9 days post the first vaccine dose timepoints; however, in the three subjects in whom both pre-vaccine and 9 days post the first vaccine dose timepoints were measured, the responses at the two timepoints were strongly correlated (r = 0.97, >0.99, and >0.99, Fig. S7A), and no trends were observed that would suggest that the observed differences in CD4+ populations between transplant and control were artifactual (Fig. S7B).

It remains unclear why natural infection is able to overcome the reduction in baseline CD4+ T cells in many transplant patients (3 of 4 in our study); moreover, in our study, two uninfected transplant patients demonstrated an antibody response, albeit delayed (Fig. 3A and 3C). Delayed responses to SARS-CoV-2 vaccination have also been reported in kidney transplant and chronic dialysis patients (20, 36). The association between IgG responses and T_PH_ cells may offer a clue, as these cells have been implicated in extrafollicular B cell differentiation and maintenance (37) and COVID-19 infection induces expansion of T_PH_ cells (38) and increases in non-germinal center B cell responses (39).

Based on data in humans and monkeys, anti-spike protein IgG and anti-RBD IgG are strong immune correlates of protection to COVID-19 infection (40–42). The weakness of this response in COVID-19-uninfected vaccinated heart and lung transplant recipients, demonstrated herein and in other studies, raises major concerns that despite vaccination, this population of patients is poorly protected from infection by standard vaccination strategies.

## Methods

### Study participants and clinical and demographic data

Eligible participants were heart and lung transplant recipients followed clinically at our institution and healthy volunteers of similar age and sex distribution. Healthy volunteers were solicited via a publicly accessible institutional website (https://rally.partners.org/) and local newsletters. Candidates for healthy controls were considered eligible if they were 30-80 years old (the same age range as enrolled transplant recipients), were not receiving immunosuppressive medications, and to enable rapid processing of obtained specimens, were living in eastern Massachusetts. All participants were enrolled prior to or up to two weeks after SARS-CoV-2 vaccination. Demographic and clinical data and vaccine type were collected from the electronic medical record and/or study participants. Evidence for prior COVID-19 infection was obtained from the electronic medical record.

### Demographics and clinical characterization of participants

Eighteen heart and eight lung transplant recipients and 12 healthy volunteers receiving SARS-CoV-2 vaccination were enrolled. The age and sex distribution of the transplant and control groups were similar (Table 1). About one half of the heart and lung transplant recipients were 1 to 5 years after transplantation, about 15% of each group were less than one year after transplantation, and one-third of heart and one-quarter of lung transplant recipients were 10 or more years after transplantation (Table 1). For the two-dose vaccines (BNT162b2 and mRNA-1273), the time between vaccine doses reflected each vaccine’s prescribed dosing schedule, with the exception of one outlier, whose second dose was delayed due to intercurrent COVID-19 infection (Fig. S6).

**Table 1.**
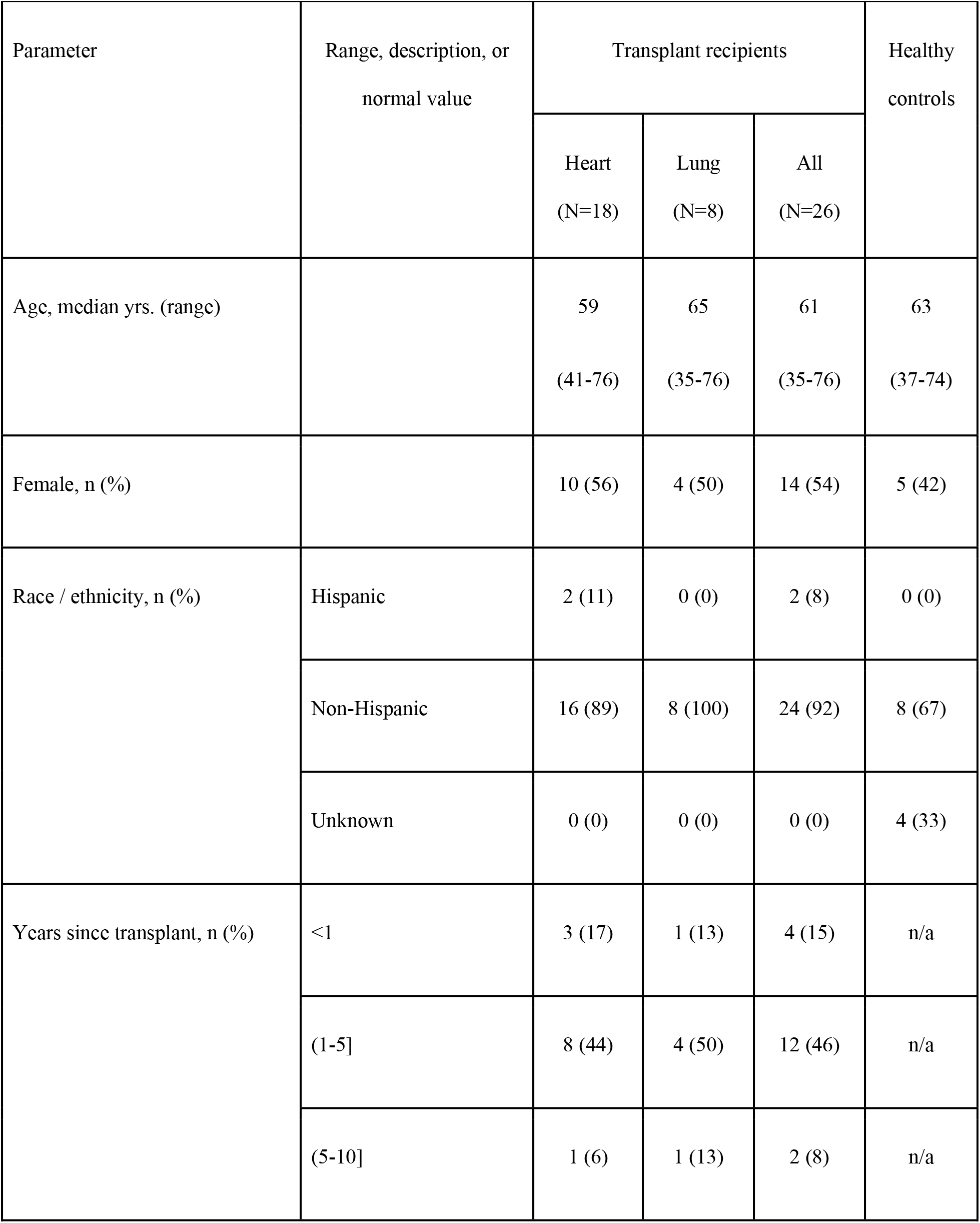
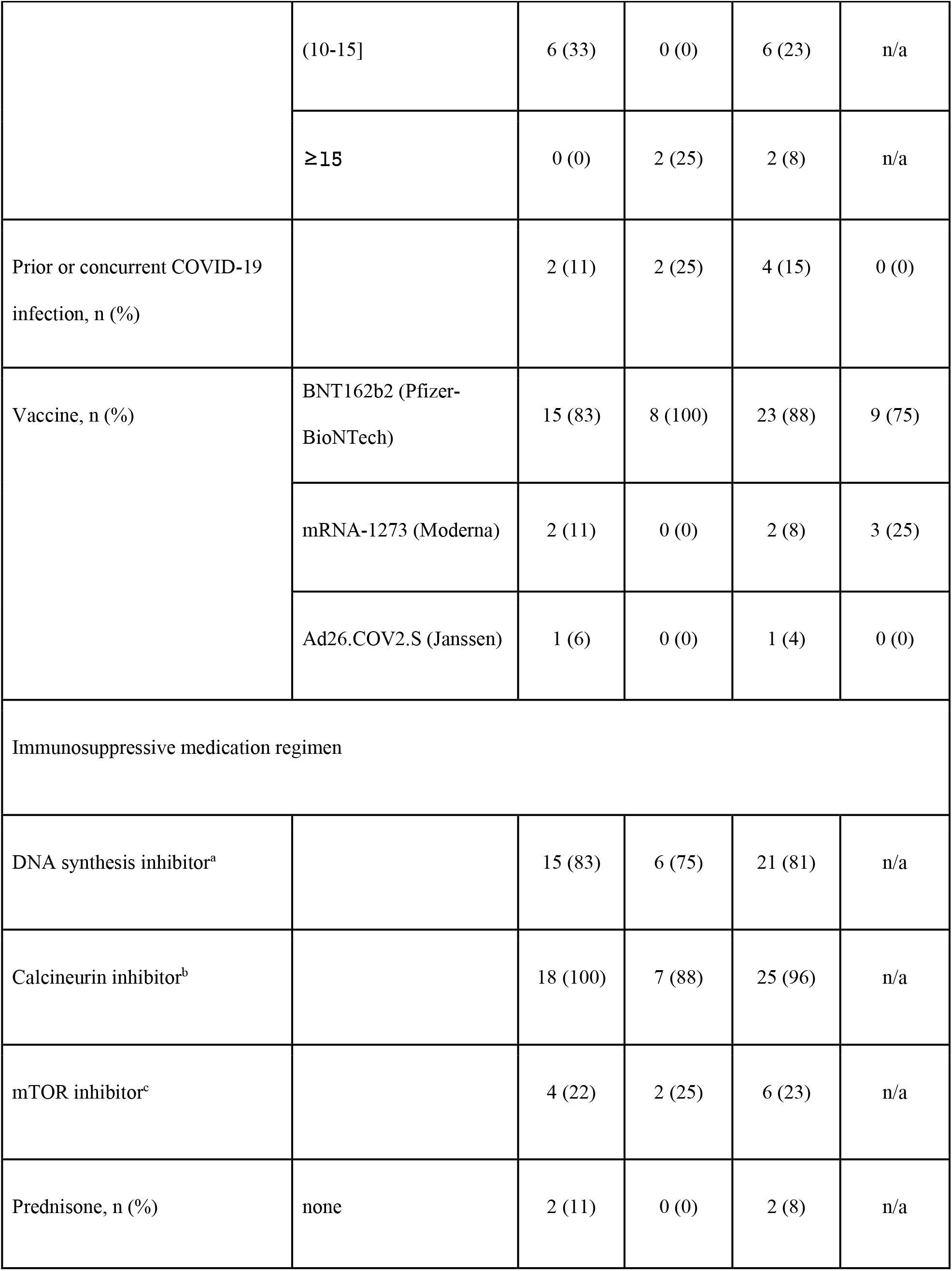
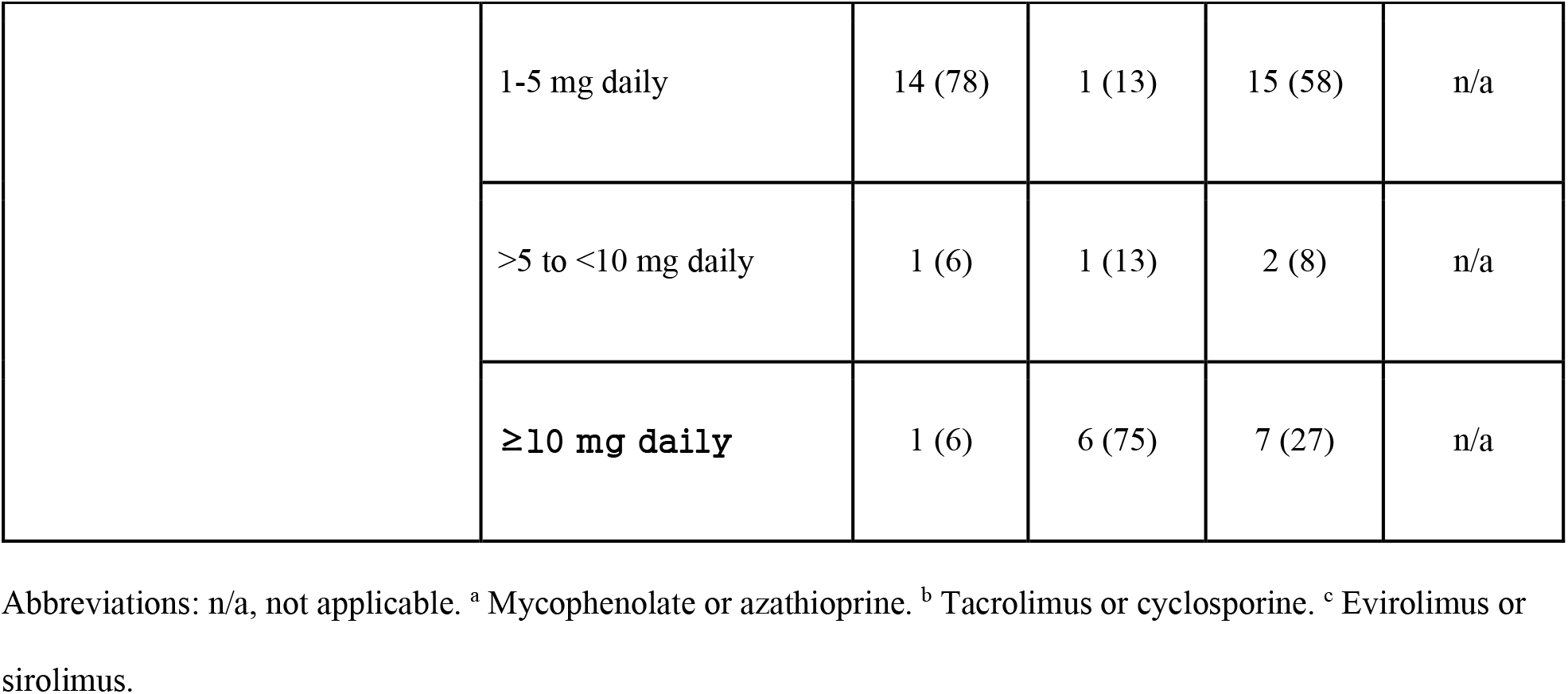
Demographic and clinical characteristics of cohort

| Parameter | Range, description, or normal value | Transplant recipients |  |  | Healthy controls |
| --- | --- | --- | --- | --- | --- |
|  |  | Heart<br>(N=18) | Lung<br>(N=8) | All<br>(N=26) |  |
| Age, median yrs. (range) |  | 59<br>(41-76) | 65<br>(35-76) | 61<br>(35-76) | 63<br>(37-74) |
| Female, n (%) |  | 10 (56) | 4 (50) | 14 (54) | 5 (42) |
| Race / ethnicity, n (%) | Hispanic | 2 (11) | 0 (0) | 2 (8) | 0 (0) |
|  | Non-Hispanic | 16 (89) | 8 (100) | 24 (92) | 8 (67) |
|  | Unknown | 0 (0) | 0 (0) | 0 (0) | 4 (33) |
| Years since transplant, n (%) | <1 | 3 (17) | 1 (13) | 4 (15) | n/a |
|  | (1-5] | 8 (44) | 4 (50) | 12 (46) | n/a |
|  | (5-10] | 1 (6) | 1 (13) | 2 (8) | n/a |
|  | (10-15] | 6 (33) | 0 (0) | 6 (23) | n/a |
|  | ≥15 | 0 (0) | 2 (25) | 2 (8) | n/a |
| Prior or concurrent COVID-19 infection, n (%) |  | 2 (11) | 2 (25) | 4 (15) | 0 (0) |
| Vaccine, n (%) | BNT162b2 (Pfizer-BioNTech) | 15 (83) | 8 (100) | 23 (88) | 9 (75) |
|  | mRNA-1273 (Moderna) | 2 (11) | 0 (0) | 2 (8) | 3 (25) |
|  | Ad26.COV2.S (Janssen) | 1 (6) | 0 (0) | 1 (4) | 0 (0) |
| Immunosuppressive medication regimen |  |  |  |  |  |
| DNA synthesis inhibitor<sup>a</sup> |  | 15 (83) | 6 (75) | 21 (81) | n/a |
| Calcineurin inhibitor<sup>b</sup> |  | 18 (100) | 7 (88) | 25 (96) | n/a |
| mTOR inhibitor<sup>c</sup> |  | 4 (22) | 2 (25) | 6 (23) | n/a |
| Prednisone, n (%) | none | 2 (11) | 0 (0) | 2 (8) | n/a |
|  | 1-5 mg daily | 14 (78) | 1 (13) | 15 (58) | n/a |
|  | >5 to <10 mg daily | 1 (6) | 1 (13) | 2 (8) | n/a |
|  | ≥10 mg daily | 1 (6) | 6 (75) | 7 (27) | n/a |

Abbreviations: n/a, not applicable. <sup>a</sup> Mycophenolate or azathioprine. <sup>b</sup> Tacrolimus or cyclosporine. <sup>c</sup> Evirolimus or sirolimus.

The vast majority of both heart and lung transplant recipient participants were maintained on a DNA synthesis inhibitor, a calcineurin inhibitor, and prednisone; about a quarter of both heart and lung transplant recipients were also maintained on an mTOR inhibitor (Table 1). Lung transplant recipients were generally maintained on higher doses of prednisone than heart transplant recipients; whereas 75% (6/8) of lung transplant recipients were taking ≥10 mg of prednisone daily, only 6% (1/18) of heart transplant recipients were taking ≥10 mg daily. No other substantial medication differences between heart transplant and lung transplant participants were observed.

### Sample collection

Serial blood samples were collected prior to vaccination (N=13), approximately 1 week after the first dose of vaccine (N=18, median 9 days, interquartile range [IQR, 25%-75%] 7-10 days), just prior to the second dose of vaccine (N=30, median 21 days, IQR 19-21 days after first dose), and at approximately 1 week (N=32, median 8 days, IQR 7-10 days), 4 weeks (N=16, median 30 days, IQR 26-34 days), and 8-10 weeks (N=18, median 60 days, IQR 60-69 days) after the second dose of vaccine. Baseline blood samples were defined as a sample collected prior to vaccination (N=12, including 4 transplant recipients and 8 healthy controls) or a blood sample collected approximately 1 week after the first dose of vaccine (N=9, all transplant recipients). Blood collection was either in the institutional Translational and Clinical Research Center or the participant’s home by Partners HealthCare at Home. All samples were processed within 4 hours of phlebotomy. Blood samples were centrifuged, and plasma, serum, and PBMCs were isolated and cryopreserved at −180°C. PBMC isolation was performed using BD cell preparation tubes with sodium heparin. Tubes were centrifuged at 1800 x g for 25 minutes. PBMC were washed in HEPES-buffered Hanks saline solution, resuspended in FBS with 10% DMSO, and stored in liquid nitrogen.

### ELISA and neutralizing antibody assays

Purified SARS-CoV-2 D614G spike protein receptor binding domain (RBD) and whole spike protein were generously provided by Aaron Schmidt. RBD enzyme linked immunosorbent assays (ELISAs) to IgG in participant serum were performed as previously described (43). Measurements were performed in triplicate and the mean of the three replicates was used. Absolute concentrations were determined based on a standard curve fitted in R using the drm package. Inferred values above 1.25mg/uL, twice the value of the most concentrated standard, were replaced with 1.25 mg/uL, a procedure which affected only two samples (9d post dose 1 and 1d pre dose 2) from one individual, a heart transplant recipient with prior COVID-19. Neutralizing activity of plasma against SARS-CoV-2 D614G, N501Y/D614G (representative of RBD of variant B.1.1.7), and E484K/N501Y/D614G (representative of RBD of variants B1.351 and P.1) spike protein pseudoviruses were assayed as previously described (44). pCMV-SARS2SΔC D614G-gp41 was generated by PCR mutagenesis from pCMV-SARS2SΔC-gp41 and Gibson assembly. pCMV-SARS2SΔC N501Y/D614G-gp41 and pCMV-SARS2SΔC E484K/N501Y/D614G-gp41 were obtained by Gibson assembly of the respective gBlocks (IDT) in pCMV-SARS2SΔC D614G-gp41.

### Flow cytometry

To comprehensively quantitate proportions of B and T cell subsets at baseline, PBMCs were studied using multicolor flow cytometry. Preserved PBMCs were thawed and washed in complete DMEM. Prior to antibody staining, Fc receptors were blocked using Human TruStain FcX (BioLegend, 422302) at a concentration of 1:20 on ice for 15 minutes. Cells were stained sequentially, first for 20 minutes at 37ºC, then washed and stained with a second cocktail of antibodies for 30 minutes at 4ºC, as previously optimized for each clone. The following antibody panel was used to quantitate B and T cell subsets: anti-human HLA-DR-BUV395 (BD Biosciences, Clone Tu39, 1:400), anti-human CD45RA-BUV563 (BD Biosciences, Clone HI100, 1:600), anti-human CD38-BUV661 (BD Biosciences, Clone HIT2, 1:400), anti-human CCR7-BUV737 (BD Biosciences, Clone 3D12, 1:200), anti-human CD4-BUV805 (BD Biosciences, Clone SK3, 1:100), anti-human CD20-BUV805 (BD Biosciences, Clone 2H7, 1:100), anti-human CD25-BV421 (BioLegend, Clone BC96, 1:25), anti-human CD28-BV480 (BD Biosciences, Clone CD28.2, 1:100), anti-human CD21-BV480 (BD Biosciences, Clone B-ly4, 1:100), anti-human CD11c-BV605 (BD Biosciences, Clone B-ly6, 1:100), anti-human CD10 (BD Biosciences, Clone HI10α, 1:25), anti-human PD1-BV711 (BD Biosciences, Clone EH12.1, 1:100), anti-human CD86-BV750 (BD Biosciences, Clone FUN-1, 1:100), anti-human CD8-BV785 (BioLegend, Clone SK1, 1:400), anti-human IgD-BV785 (BioLegend, Clone 1A6-2, 1:100), anti-human CXCR5-A488 (BD Biosciences, Clone RF8B2, 1:100), anti-human CD127-BB700 (BD Biosciences, Clone HIL-7R-M21, 1:50), anti-human CX3CR1-PE (BioLegend, Clone K0124E1, 1:25), anti-human CD27-PE-Dazzle (BioLegend, Clone M-T271, 1:100), anti-human CXCR3-PE-Cy7 (BioLegend, Clone G025H7, 1:50), anti-human CCR6-APC (BioLegend, Clone G034E3, 1:25), anti-human CD19-APC-R700 (BD Biosciences, Clone SJ25C1, 1:25), and anti-human CD3-APC-Vio770 (Milltenyi, Clone BW264/56, 1:200).

After staining, cells were washed with 1% BSA in PBS, centrifuged and resuspended in 1% BSA in PBS. Just prior to flow cytometry, dead cells were identified with SYTOX AADvanced Dead Cell Stain (Thermo Fisher Scientific, S10274, 1:1000). Flow cytometry was performed on a BD FACS Symphony cytometer (BD Biosciences, San Jose, CA), and rainbow tracking beads were used to ensure consistent signals between flow cytometry batches. FCS files were analyzed using FlowJo software (version 10.7.2). The surface markers used to define each cell type are provided in Table 2.

**Table 2.**
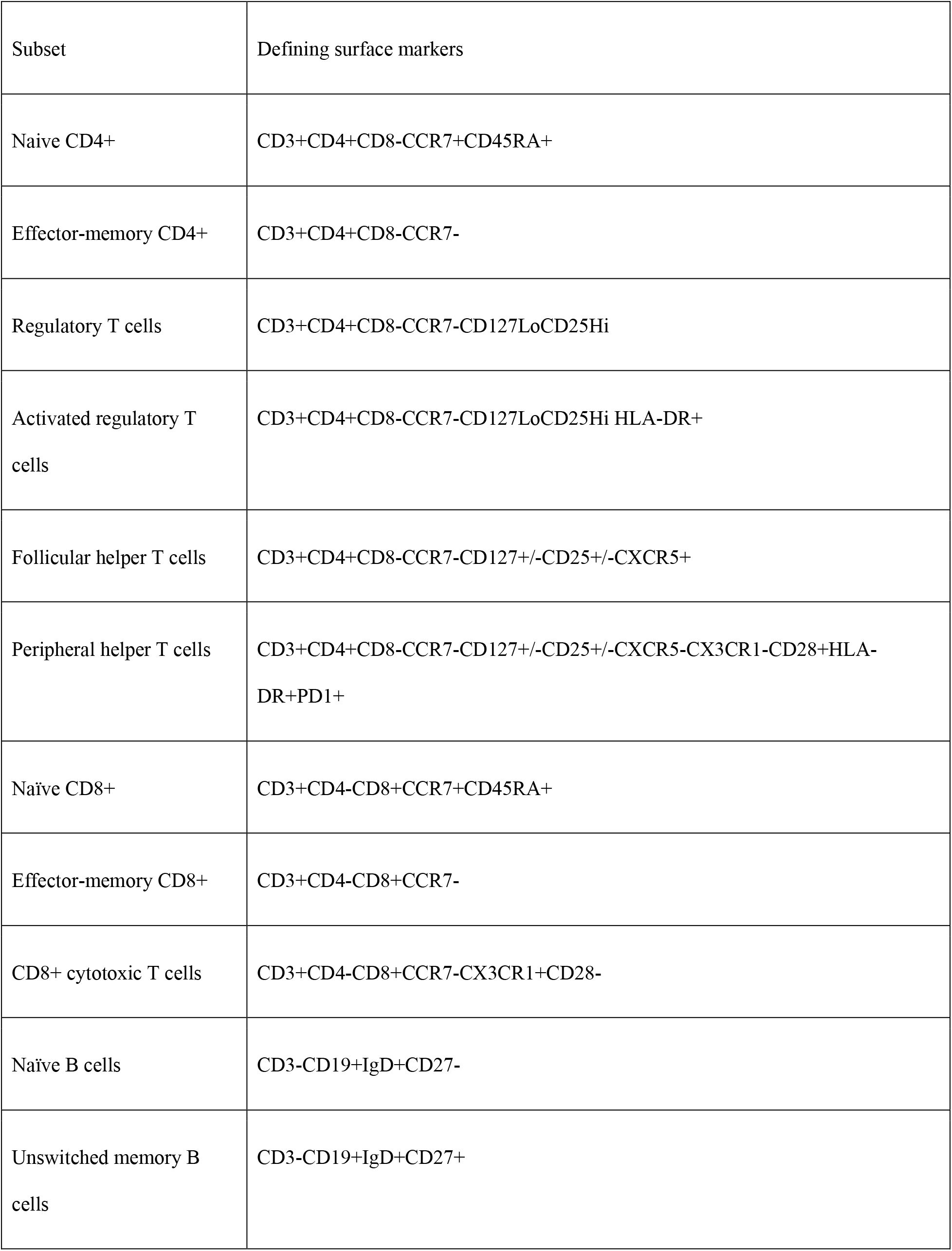
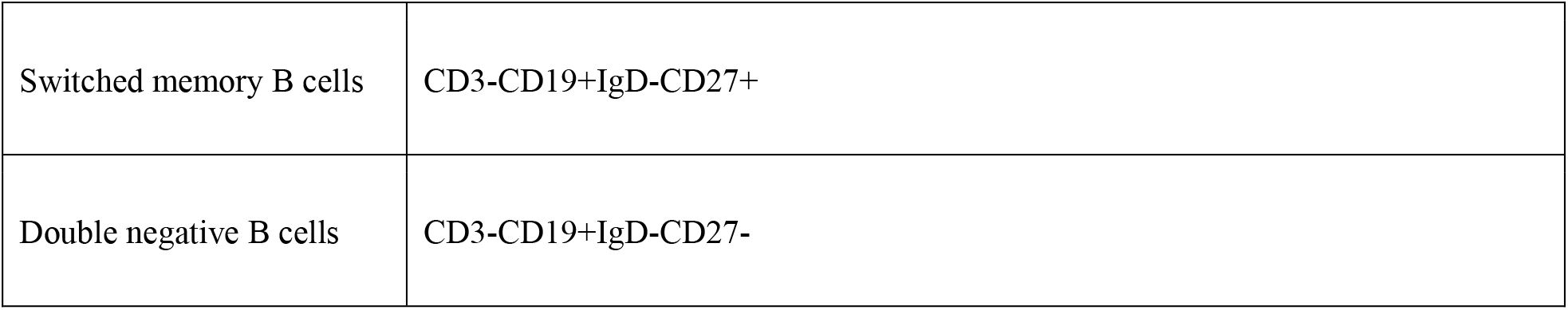
Surface markers used to define T and B cell subsets

| Subset | Defining surface markers |
| --- | --- |
| Naïve CD4+ | CD3+CD4+CD8-CCR7+CD45RA+ |
| Effector-memory CD4+ | CD3+CD4+CD8-CCR7- |
| Regulatory T cells | CD3+CD4+CD8-CCR7-CD127LoCD25Hi |
| Activated regulatory T cells | CD3+CD4+CD8-CCR7-CD127LoCD25Hi HLA-DR+ |
| Follicular helper T cells | CD3+CD4+CD8-CCR7-CD127+/-CD25+/-CXCR5+ |
| Peripheral helper T cells | CD3+CD4+CD8-CCR7-CD127+/-CD25+/-CXCR5-CX3CR1-CD28+HLA-DR+PD1+ |
| Naïve CD8+ | CD3+CD4-CD8+CCR7+CD45RA+ |
| Effector-memory CD8+ | CD3+CD4-CD8+CCR7- |
| CD8+ cytotoxic T cells | CD3+CD4-CD8+CCR7-CX3CR1+CD28- |
| Naïve B cells | CD3-CD19+IgD+CD27- |
| Unswitched memory B cells | CD3-CD19+IgD+CD27+ |
| Switched memory B cells | CD3-CD19+IgD-CD27+ |
| Double negative B cells | CD3-CD19+IgD-CD27- |

A single aliquot of PBMC was thawed and used for flow cytometry. Each aliquot corresponds to cells harvested from a single 8 mL CPT; however, absolute counts may reflect additional noise incorporated during a single cycle of freezing and thawing. We therefore also report relative counts as a proportion of CD4+, CD8+ and CD19+ cells.

### Statistics

Raw data were analyzed with R (45). Pairwise comparisons were performed using a Wilcoxon test. Correlation coefficients and associated p-values were calculated using Pearson’s product-moment correlation coefficient. The proportion of subjects who responded to vaccination was calculated by Fisher’s exact test. Response to vaccination for each subject was determined by a consistent rise in IgG ELISA over time. Data were displayed using ggplot (46).

### Study approval

All study procedures involving human subjects were approved by the Mass General Brigham Human Research Committee, the governing institutional review board at Massachusetts General Hospital. Informed consent was received from participants prior to inclusion in the study, either in writing or by institutional review board-established verbal consent procedures employed during the COVID-19 pandemic. Participants were identified by numbers.

## Data Availability

All data are included in the manuscript and/or will be made available upon request.

## Author contributions

J.E.L., designed research studies, recruited and consented patients, scheduled patients, conducted experiments, acquired data, analyzed data, wrote manuscript.

A.L., designed research studies, analyzed data, wrote manuscript.

M.G., conducted experiments, acquired data, edited manuscript.

C.A.P., designed research studies, conducted experiments, acquired data, edited manuscript.

Z.W., recruited and consented patients, scheduled patients, acquired data, edited manuscript.

K.B., recruited and consented patients, scheduled patients, edited manuscript.

P.A., recruited and consented patients, scheduled patients, edited manuscript.

H.L., conducted experiments, acquired data.

G.D.L., recruited patients, edited manuscript.

N.B., conducted experiments, acquired data.

T.L., recruited and consented patients, scheduled patients.

N.H., designed research studies.

S.S.P., designed research studies, analyzed data, edited manuscript.

M.B.G., designed research studies, recruited and consented patients, scheduled patients, analyzed data, wrote manuscript, raised funds to support studies.

## Acknowledgments

This work was supported by the American Lung Association COVID-19 Action Initiative Research Award (to M.B.G.), the Cystic Fibrosis Foundation (to M.B.G.), Doris Duke Charitable Foundation Physician-Scientist Fellowship (to J.E.L.), a CRI/Bristol Myers Squibb Postdoctoral Fellowship (CRI 2993) (to M.G.), NIH U19 AI 110495 (to S.S.P.), and NIH UL1 TR 002541-01 (to the Massachusetts General Hospital Translational and Clinical Research Center).

We are grateful to Grace Holland and Kathy Hall of the Massachusetts General Hospital Translational and Clinical Research Center, Gary Garberg and Jennifer Erickson of Partners HealthCare at Home, Meaghan Doucette, Aaron Schmidt, and Richelle Charles.

