## Supplemental Figures for "Vaccine serologic responses among transplant patients associate with COVID-19 infection and T peripheral helper cells"

**Jacob E. Lemieux, et al.**

### Supplemental Figures

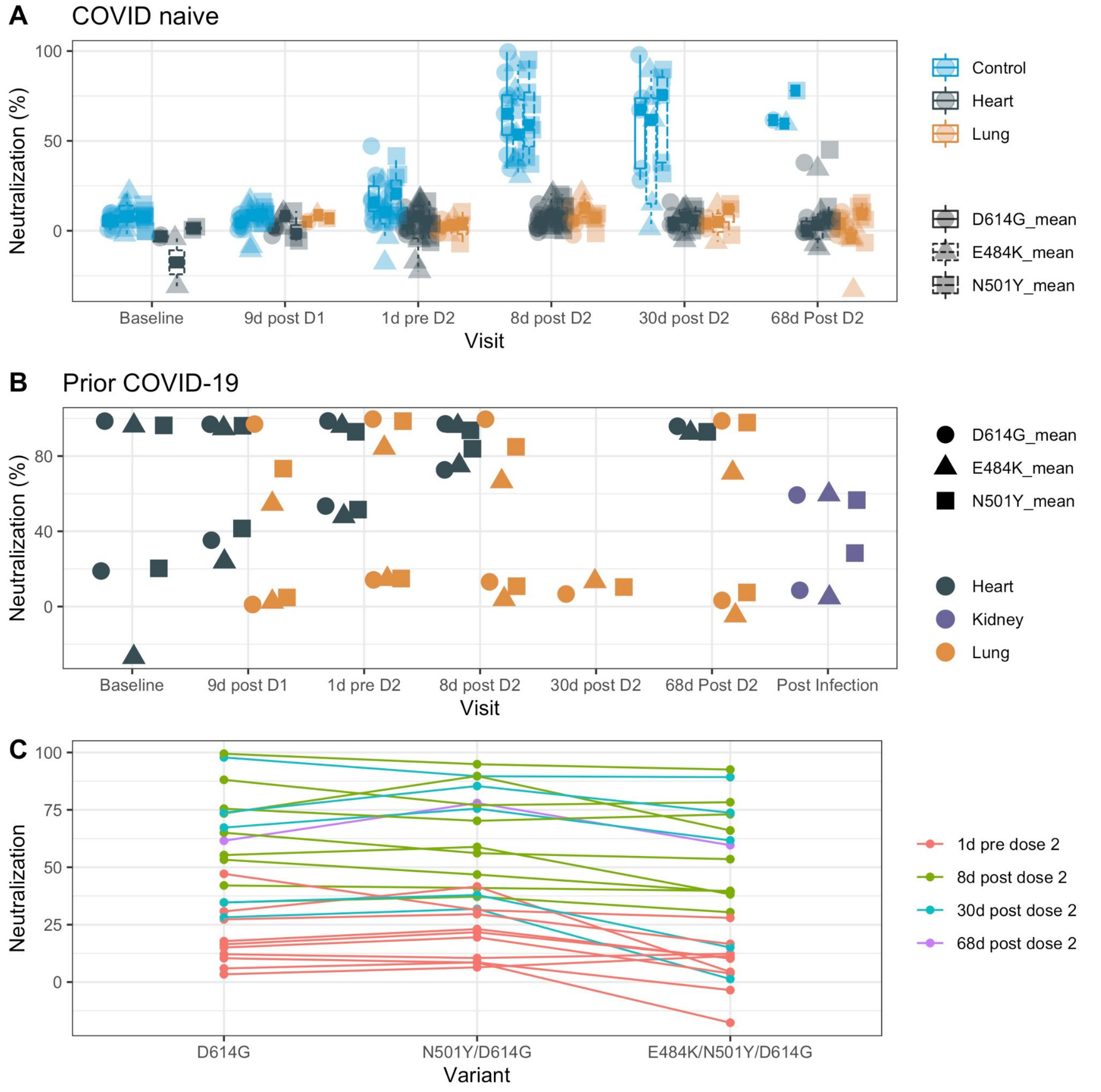


**Supplemental Figure 1: Neutralization activity against pseudovirus variants.** (**A**) Neutralization activity against the indicated pseudovirus variants for COVID-19-naive subjects (**A**) or subjects with prior COVID-19 infection (**B**) displayed by transplanted organ type. D614G, D614G variant pseudovirus; E484K, E484K/N501Y/D614G pseudovirus; N501Y, N501Y/D614G pseudovirus. For the analysis in (**A**), boxes depict the 25$^{th}$, 50$^{th}$, and 75$^{th}$ percentiles; whiskers, smallest and largest values in dataset up to 1.5x interquartile range. Values lying outside whiskers are plotted individually. All data points are shown. Due to the small number of data points in (**B**), boxes and whiskers were not displayed. (**C**) Neutralization activity by variant among samples from healthy controls, colored by time post vaccination.

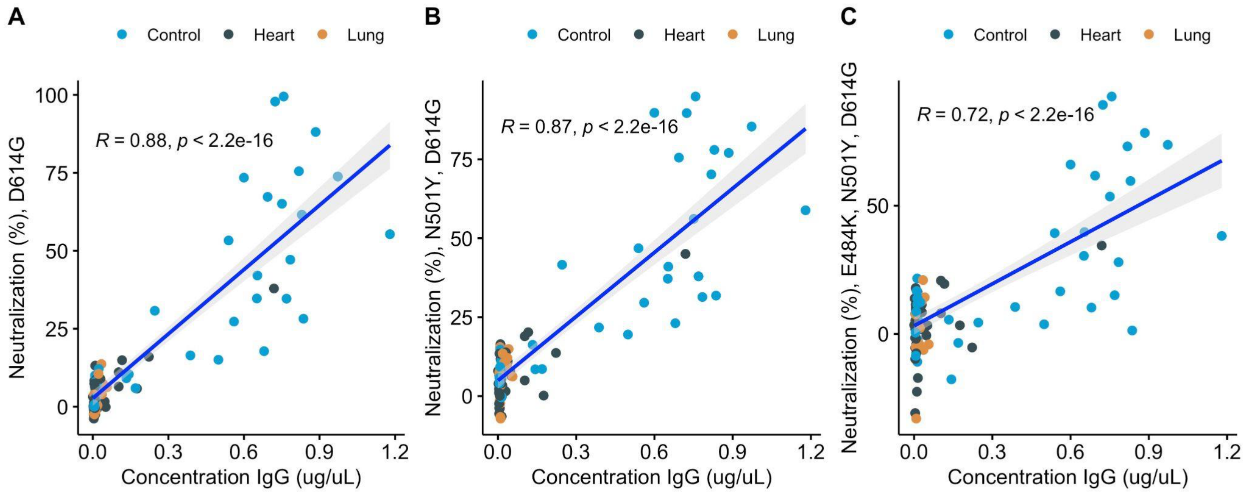


**Supplemental Figure 2: Correlation of ELISA and pseudovirus neutralization activity by pseudovirus variant.** Correlation of neutralization activity against variant pseudoviruses to anti-RBD IgG levels, displayed by transplanted organ type. Variant pseudovirus D614G (**A**), N501Y/D614G (**B**), and E484K/N501Y/D614G (**C**). Least squares regressions (blue lines) of neutralization activity to indicated variant pseudoviruses. Gray shading, 95% confidence interval (CI). *R*, correlation coefficient. The plotted data points include all available time points (pre dose 1 through 68d post dose 2) from both subjects who are COVID-19 naive and who had prior or intercurrent COVID-19.

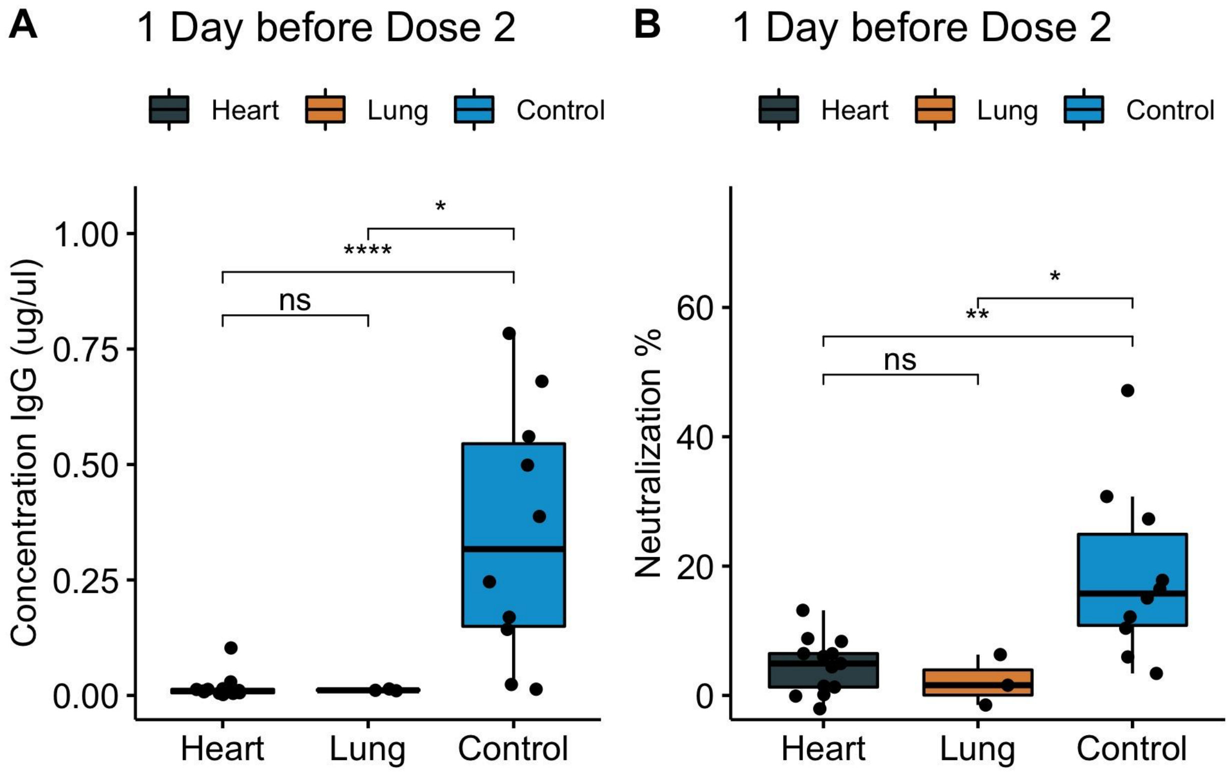


**Supplemental Figure 3: Levels of serum IgG to SARS-CoV-2 RBD and pseudovirus neutralization activity just prior to the second dose of vaccine among COVID-19 naive subjects.** (**A**) Anti-spike RBD IgG binding 1 day before the second vaccine dose, displayed by transplanted organ type. Measured by ELISA. (**B**) SARS-CoV-2 pseudovirus neutralization activity, 1 day before the second dose, displayed by transplanted organ type. Data for the transplant recipients who had prior COVID-19 infection are excluded. Boxes, $25^{th}$, $50^{th}$, and $75^{th}$ percentiles; whiskers, smallest and largest value in dataset up to 1.5X interquartile range. All data points are shown. ns, not significant; *, $p < 0.05$; **, $p < 0.01$; ****, $p < 0.0001$.

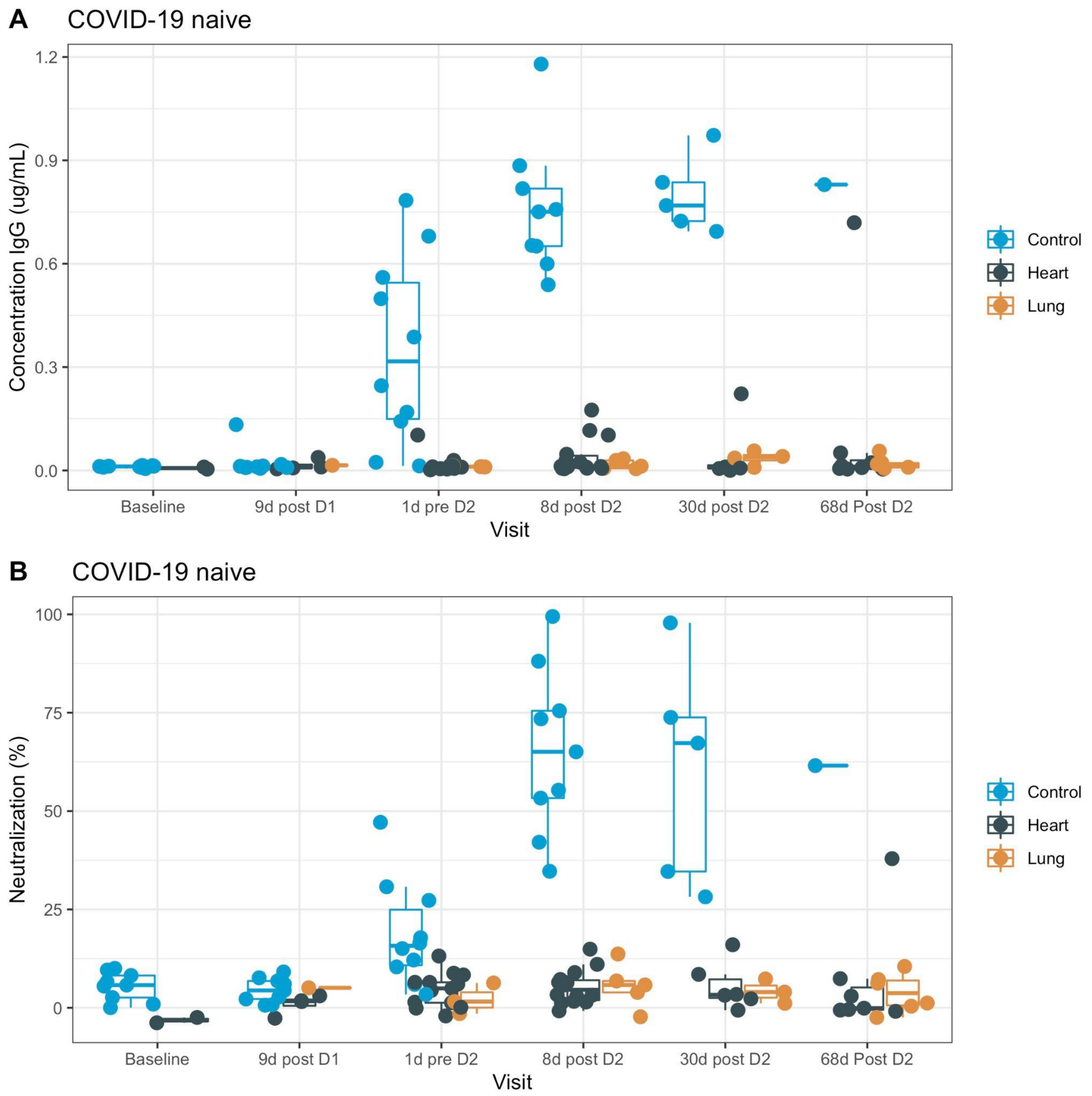


**Supplemental Figure 4: Time course of IgG antibody responses among COVID-19 naive subjects.** (**A**) IgG binding to SARS-CoV-2 spike RBD, as measured by ELISA, at each of the first 6 study timepoints. (**B**) SARS-CoV-2 neutralization at each of the first 6 study timepoints. Data displayed by transplanted organ type. Boxes, 25th, 50th, and 75th percentile; whiskers, smallest and largest values in dataset up to 1.5x interquartile range. All data points are shown.

**A**

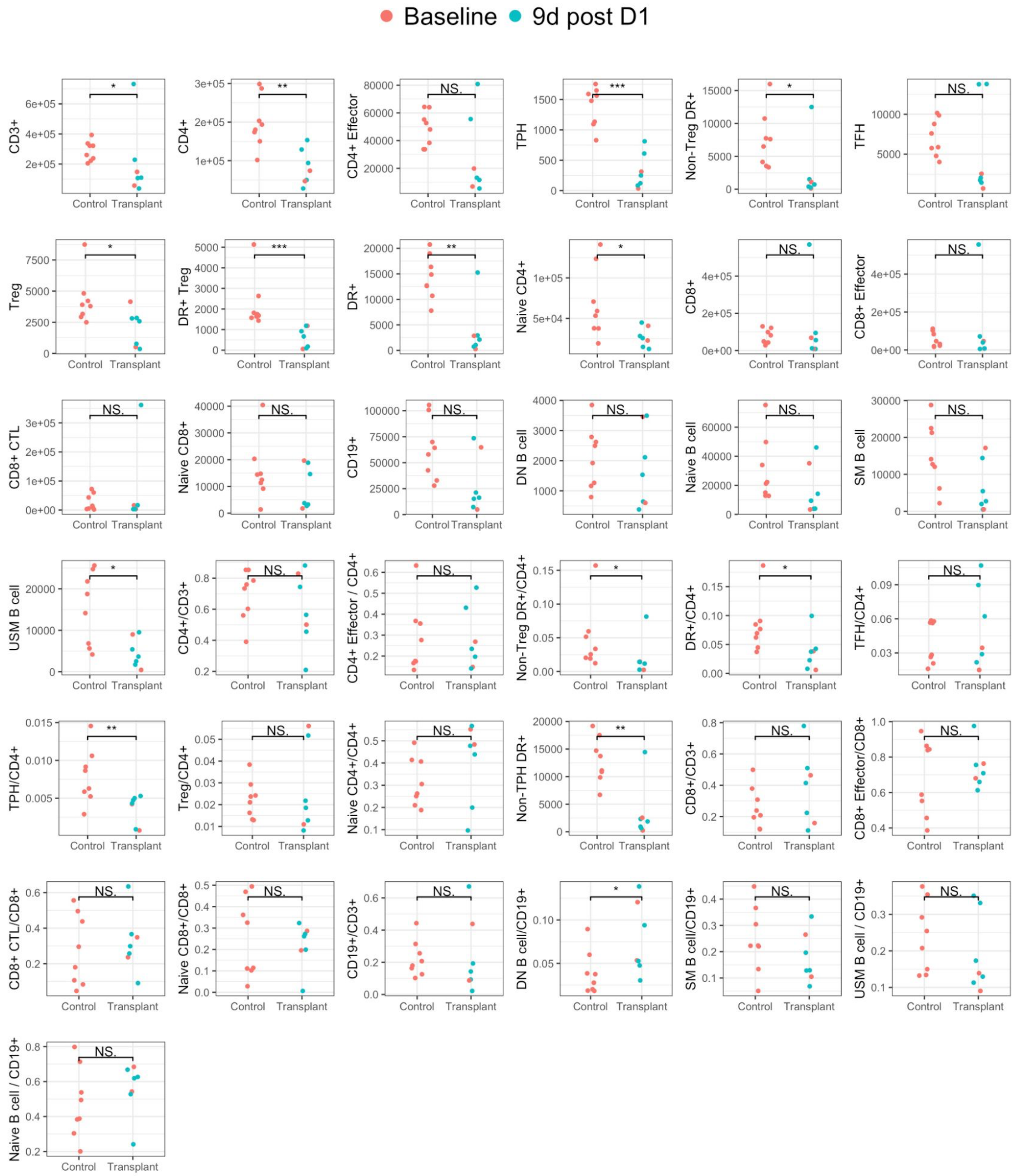
Baseline
9d post D1
CD3+
CD4+
CD4+ Effector
TPH
Non-Treg DR+
TFH
Treg
DR+ Treg
DR+
Naive CD4+
CD8+
CD8+ Effector
CD8+ CTL
Naive CD8+
CD19+
DN B cell
Naive B cell
SM B cell
USM B cell
CD4+/CD3+
CD4+ Effector / CD4+
Non-Treg DR+/CD4+
DR+/CD4+
TFH/CD4+
TPH/CD4+
Treg/CD4+
Naive CD4+/CD4+
Non-TPH DR+
CD8+/CD3+
CD8+ Effector/CD8+
CD8+ CTL/CD8+
Naive CD8+/CD8+
CD19+/CD3+
DN B cell/CD19+
SM B cell/CD19+
USM B cell / CD19+
Naive B cell / CD19+
Control
Transplant
NS.
*
**
***

**B**

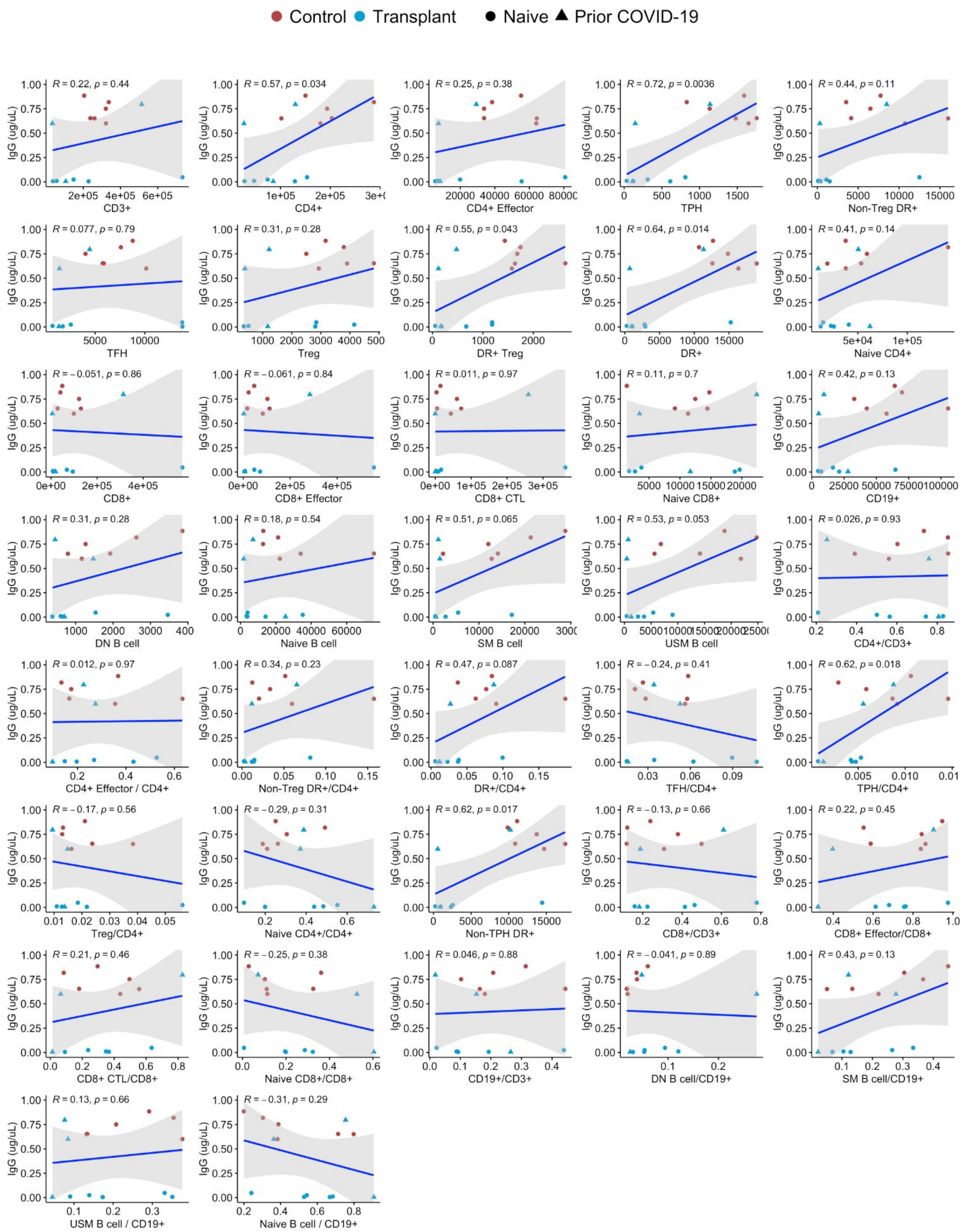

Control Transplant Naive Prior COVID-19
IgG (ug/uL)
R = 0.22, p = 0.44
CD3+
R = 0.57, p = 0.034
CD4+
R = 0.25, p = 0.38
CD4+ Effector
R = 0.72, p = 0.0036
TPH
R = 0.44, p = 0.11
Non-Treg DR+
R = 0.077, p = 0.79
TFH
R = 0.31, p = 0.28
Treg
R = 0.55, p = 0.043
DR+ Treg
R = 0.64, p = 0.014
DR+
R = 0.41, p = 0.14
Naive CD4+
R = −0.051, p = 0.86
CD8+
R = −0.061, p = 0.84
CD8+ Effector
R = 0.011, p = 0.97
CD8+ CTL
R = 0.11, p = 0.7
Naive CD8+
R = 0.42, p = 0.13
CD19+
R = 0.31, p = 0.28
DN B cell
R = 0.18, p = 0.54
Naive B cell
R = 0.51, p = 0.065
SM B cell
R = 0.53, p = 0.053
USM B cell
R = 0.026, p = 0.93
CD4+/CD3+
R = 0.012, p = 0.97
CD4+ Effector / CD4+
R = 0.34, p = 0.23
Non-Treg DR+/CD4+
R = 0.47, p = 0.087
DR+/CD4+
R = −0.24, p = 0.41
TFH/CD4+
R = 0.62, p = 0.018
TPH/CD4+
R = −0.17, p = 0.56
Treg/CD4+
R = −0.29, p = 0.31
Naive CD4+/CD4+
R = 0.62, p = 0.017
Non-TPH DR+
R = −0.13, p = 0.66
CD8+/CD3+
R = 0.22, p = 0.45
CD8+ Effector/CD8+
R = 0.21, p = 0.46
CD8+ CTL/CD8+
R = −0.25, p = 0.38
Naive CD8+/CD8+
R = 0.046, p = 0.88
CD19+/CD3+
R = −0.041, p = 0.89
DN B cell/CD19+
R = 0.43, p = 0.13
SM B cell/CD19+
R = 0.13, p = 0.66
USM B cell / CD19+
R = −0.31, p = 0.29
Naive B cell / CD19+

**Supplemental Figure 5: Immune subsets in peripheral blood at baseline and correlation of baseline flow cytometry with IgG responses at 8 days after the 2nd vaccine dose.** (**A**) Quantification by flow cytometry of peripheral blood cells in COVID-19 naive individuals. All data points for COVID-19 naive individuals are shown. *, $p < 0.05$; **, $p < 0.01$; ***, $p < 0.001$; ****, $p < 0.0001$. (**B**) Scatterplots of IgG response vs. baseline flow cytometry. Least squares regressions (blue lines) of IgG responses to the indicated cell numbers or ratios, as indicated. Gray shading, 95% confidence interval (CI). The earliest available sample for each patient (from pre dose 1 or day 8 post-dose 1) is shown; patients with prior COVID-19 are denoted by triangles. *R*, correlation coefficient. Treg, regulatory T cells; TFH, T follicular helper cells; CTL, cytotoxic T cell; DN B cells, double negative B cells; TPH, T peripheral helper cells; CD4 Effector, effector-memory CD4+ T cells; CD8 Effector, effector-memory CD8+ T cells; SM B cells, switched memory B cells; USM B cells, unswitched memory B cells.

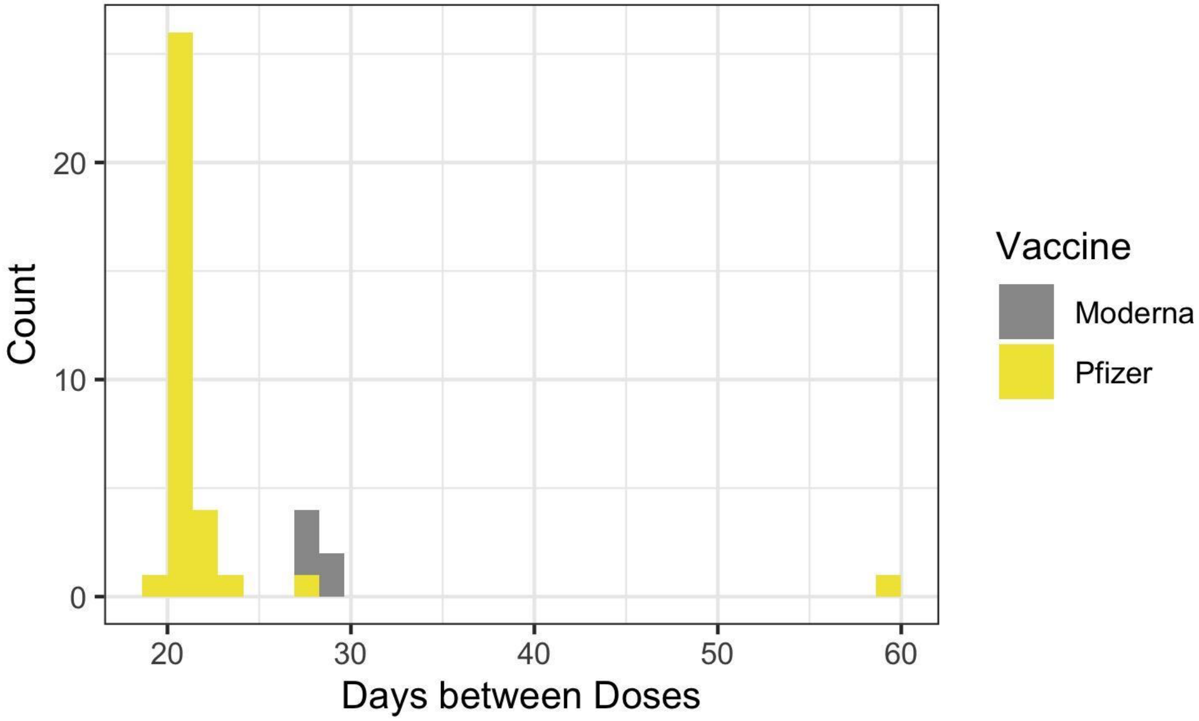


**Supplemental Figure 6: Time between doses by vaccine type.** Distribution of time interval between doses of BNT162b2 (Pfizer-BioNTech) mRNA vaccine and mRNA-1273 (Moderna) vaccine among study participants. A single participant who received Ad26.COV2.S (Janssen) is not shown because no interval between doses could be calculated for a single dose.

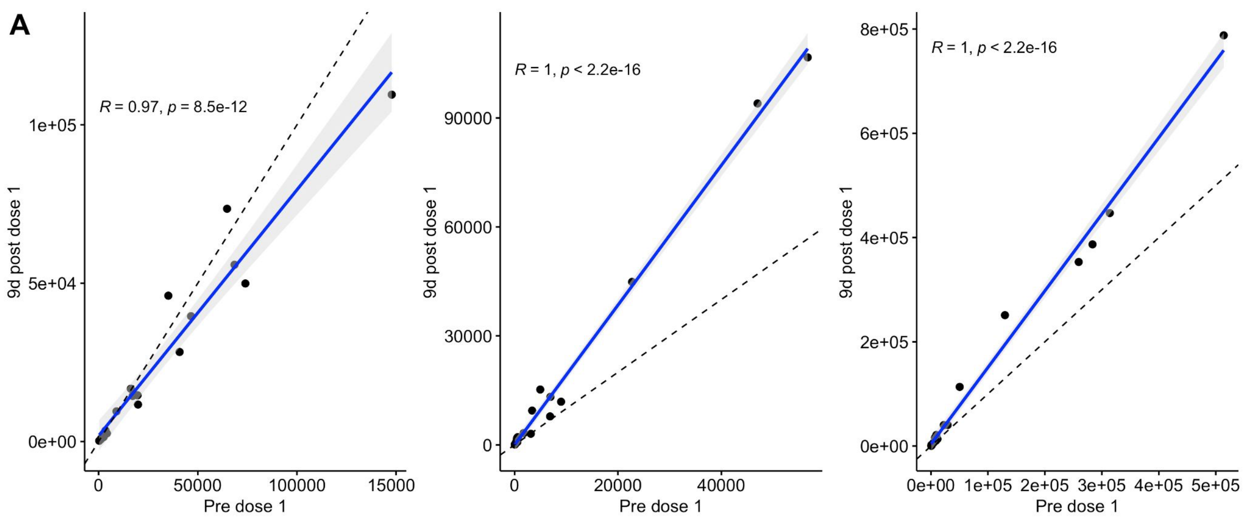
A
R = 0.97, p = 8.5e-12
1e+05
5e+04
0e+00
9d post dose 1
0
50000
100000
15000
Pre dose 1
R = 1, p < 2.2e-16
90000
60000
30000
0
9d post dose 1
0
20000
40000
Pre dose 1
R = 1, p < 2.2e-16
8e+05
6e+05
4e+05
2e+05
0e+00
9d post dose 1
0e+00
1e+05
2e+05
3e+05
4e+05
5e+05
Pre dose 1

**B**

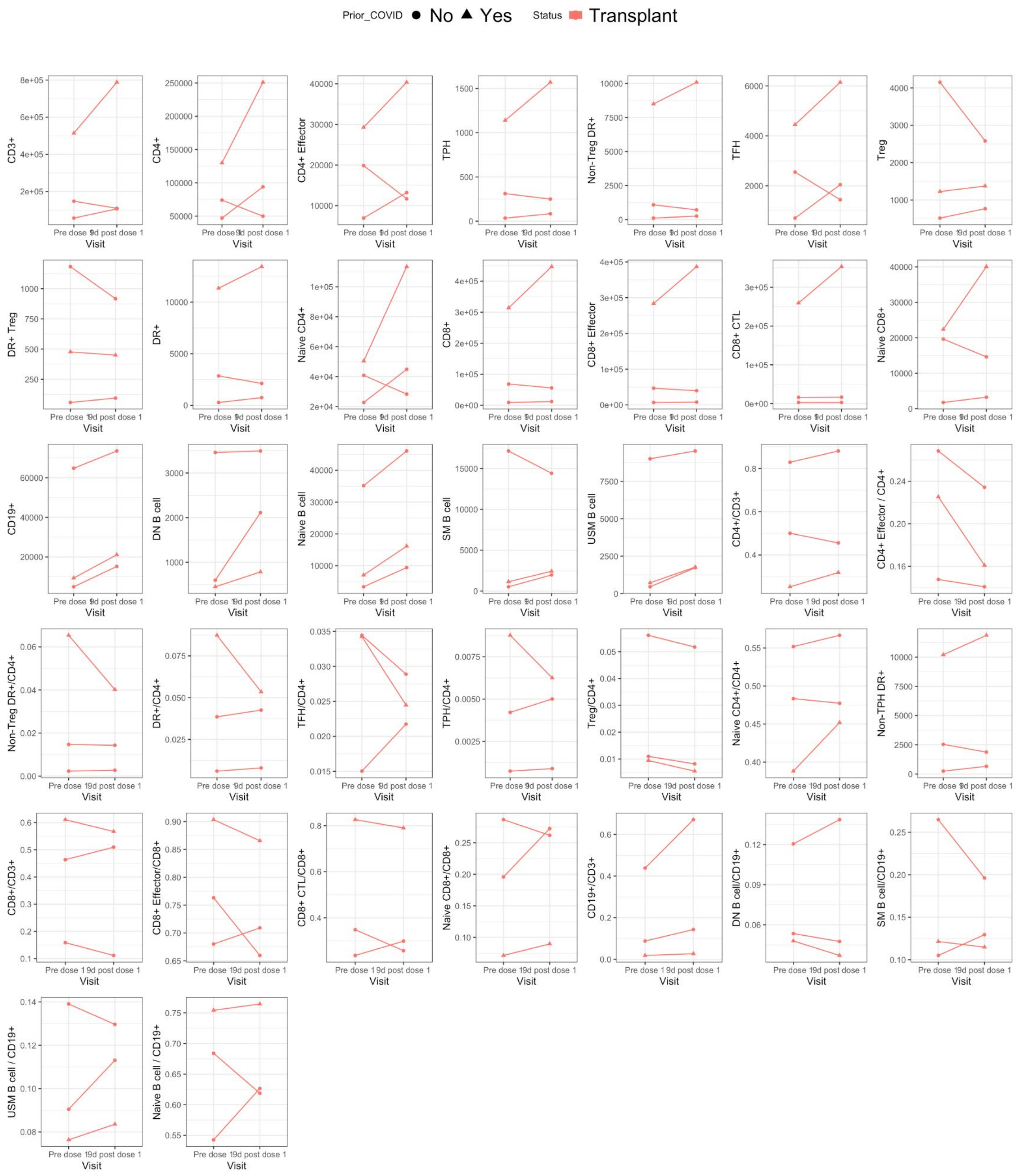


**Figure S7: Matched pre and post vaccine dose 1 cellular responses in a subset of transplant recipients.** Flow cytometry analysis of samples collected pre vaccine dose 1 and at the 9 days post vaccine dose 1 for one transplant recipient with prior COVID-19 and two COVID-19-naive transplant recipients. (**A**) Scatterplot of responses. The x-axis depicts absolute counts from flow cytometry prior to dose 1, and the y-axis depicts absolute counts from the 9d post dose 1 visit. Each point is 1 of 19 cellular populations, assessed by flow cytometry. Solid line, a least squares regression fit. Light blue shading, 95% confidence interval (CI). Dotted line, a line of identity (slope = 1). (**B**) Paired comparisons of peripheral blood immune cell analysis by flow-cytometry. T and B cell subset counts for two timepoints (pre dose 1 and 9d post dose 1) are shown, and samples from a given patient are connected by a line. Triangles, data from one transplant recipient with prior COVID-19; circles, data from two COVID-19-naive transplant recipients.
